# Biological embedding of the pyscho-social environment; an Epigenetic Analysis of Adversity from Early-life to Adulthood

**DOI:** 10.64898/2026.02.13.26345039

**Authors:** Megan Buchanan, Jeanne Le Cléac’h, Sophie B. Mériaux, Jonathan D. Turner, Archibold Mposhi

## Abstract

**Introduction:** Research has shown that social and physical stressors of early-life adversity (ELA) can negatively affect long-term health trajectories. Despite differences in types of ELA exposure, previous studies have identified common health-related outcomes in adults who had experienced less favourable conditions during developmentally sensitive periods. This meta-analysis investigates the potential role of DNA methylation in mediating these adverse health trajectories by identifying common biological signatures across cohorts with distinct adversity exposures and environmental backgrounds.

**Materials and Methods:** DNA methylation data from previously published studies was used to perform a meta-analysis on 227 individuals across three cohorts. These include the EpiPath cohort consisting of adults who were exposed early institutional care, ImmunoTwin cohort consisting of adversity discordant monozygotic twin pairs and lastly a cohort of young children exposed to early institutional care.

**Results:** DNA methylation analysis across the three cohorts revealed differential methylation at CpG loci associated with 15 genes common to all cohorts. These genes are involved in neuronal development, chromatin remodeling and metabolism. Pathway enrichment analysis of the combined dataset showed potential associations with oxytocin signalling, regulation of nervous system development, and calcium signalling in relation to the later-life phenotype of the adversity exposed individuals. In addition, a poly-epigenetic score was developed by identifying a subset of 200 differentially methylated CpG sites through PLS-DA analysis with the combined beta matrix of these cohorts.

**Conclusion:** This study highlights the long-term impact of adversity by identifying common DNA methylation signatures of negative life experiences across three cohorts. The development of a poly-epigenetic score represents the first steps towards identifying group differences by combining weighted methylation values for CpG sites of interest. This method illustrates the potential to track changes in individuals across long-term studies that may benefit research in lifelong healthoutcomes.

## Introduction

Sensitive periods in childhood development highlight the possible risks to poor long-term health outcomesafter early stress exposure. Adverse social and environmental conditions during the first 1,000 days of life, known as the Barker window, have been associated with greater susceptibility to later-life diseases [1]. From conception onwards, individual experiences of early-life adversity (ELA) can encompass different external hardships involving psychosocial, financial and physical factors [2]. In particular, early maternal separation and institutionalisation are considered severe forms of adversity as the exposure to non-continuous care is associated with behavioural and developmental issues such as inattention and impulsivity even after adoption [3]. Additional negative long-term health outcomes linked to ELA exposure include higher risks of developing cardiovascular disease [4], psychopathologies related to anxio-depressive behaviours [5], and metabolic diseases like type 2 diabetes [6]. Developmental epigenetic modifications potentially play a role in these stress-related disorders as such changes continue to be observed in adolescence following early-life stressors [7].

However, plasticity in stress response systems has been observed during certain sensitive periods depending on the environmental conditions. A study found that children placed in foster care before 24 months of age showed normalisation of their autonomic nervous system and Hypothalomic-Pituitary-Adrenal (HPA) axis functions in comparison to those who remained in institutional care [8]. In addition, the Gunnar window is a potential recalibration period during adolescence where biological systems may exhibit more typical activity levels in a newly supportive environment [9]. Recently, HPA axis reactivity to psychosocial stressors in post-institutionalised children has been shown to reach similar patterns to non-adopted peers in later puberty despite earlier observations of blunted activity [10].

The “three-hit” model of the Developmental Origins of Health (DoHAD) posits that latent phenotypes for adulthood diseases may be produced by interactions between an individual’s genome and early-life exposome [11]. This time-sensitive approach to understanding stress exposure views early developmental windows as adjustment periods for the immune system to its experienced environment [12]. Individuals may be placed on developmental trajectories that either match or mismatch their future environmental conditions whereby mismatches could increase disease risk [13]. Epigenetic changes in DNA methylation are a potential mechanism for such adjustments as alterations in gene expression of ELA individuals have been observed in pathways involved in stress regulation and HPA axis function [14].

Recent theoretical advances propose that biological embedding of adversity may reflect an active cost mechanism rather than passive damage. The “adaptive cost hypothesis” posits that environmental stressors trigger adaptive reprogramming of physiological systems to maintain vigilance and mobilize resources for potential threats[15]. This recalibration, while potentially adaptive in threatening environments, incurs ongoing metabolic and physiological costs that manifest as altered disease risk trajectories. Epigenetically, this framework predicts hypomethylation of genes involved in threat detection, immune activation, and stress responsivity—creating a “primed” biological state. Distinguishing adaptive cost from wear-and-tear models has important implications: if adversity-associated methylation changes reflect adaptive tuning rather than damage, interventions may need to focus on recalibration rather than repair.

In order to explore the “adaptive cost hypothesis” and to place it correctly within the three-hit model of the DOHaD, this study will examine three cohorts with differences in their type of adversity exposure and participant composition. Firstly, the EpiPath [16] cohort includes participants who have experienced early institutional care before being adopted. The Naumova et al. study [17] cohort consists of children institutionalised in orphanages. Lastly, the ImmunoTwin cohort incorporates adversity discordant monozygotic twins. The cohorts are well suited to explore biological effects of adversity in both the Barker and Gunnar windows due to these age and timing of exposure differences.

Here, we aim to investigate how psychosocial adversity influences DNA methylation patterns across three cohorts of early life adversity. In this study, we explore how specific DNA methylation changes are associated with psychosocial adversity at different windows of development and underline its potential influence on long-term health trajectories.

## Materials and Methods

### Human Cohorts

The study incorporated data and samples from three cohorts, including our two previously published EpiPath and ImmunoTwin cohorts[18, 19]. EpiPath consists of 115 young adults who were recruited from Luxembourg and the greater region between July 2014 and March 2016 [20]. The current study used remaining DNA sample from 111 participants with a control group of 71 individuals who lived with their biological parents. The adversity exposed group was comprised of 40 participants who had previously experienced parental separation and adoption at an early age. The majority of adversity exposed individuals had also been institutionalised before the median adoption age of 4.3 months [21].

The ImmunoTwin cohort was comprised of 29 adversity discordant monozygotic twin pairs who had a divergent interpretation of their psycho-social environment. This cohort is part of the larger German twin family study, TwinLife, which focuses on social inequality and its underlying psychosocial mechanisms.

The third cohort by Naumova et al. consists of 58 children from the Saint-Petersburg region in Russia between the ages of 8 and 35 months [17]. The biological family care group (BFC) included 29 children from low or low-middle socioeconomic backgrounds without records of neglect or child abuse. The institutional care group (IC) was comprised of 29 children from three well-equipped state-run orphanages with acceptable numbers of caregivers to provide basic physical and medical needs. The majority of IC group children underwent parental separation after birth or within one year of birth.

### Ethical Approval

Ethical approval for the EpiPath study was received by the National Research Ethics Committee of Luxembourg (CNER, reference 201303/10 v1.4) and the University of Luxembourg Ethics Review Panel (ERP, No 13–002). Written informed consent was provided by all participants who also received a small financial compensation. The TwinLife study for the ImmunoTwin cohort received ethical approval from the Ethical Committee of Bielefeld University (no. 2020-184-W1).This study also incorporated data from the previously published study by Naumova et. al and was obtained upon permission by the researchers.

### Data Analysis

Data from the three cohorts were uploaded as .idat files into GenomeStudio (Illumina) for quality control assessment and imported into R Studio (R version 4.4.2) for recommended pre-processing with SeSAMe pipeline (Sensible step-wise analysis of DNA methylation bead chips). The readIDATpair() function was used to import raw IDAT files as sdf objects into the R workspace using the “EPICv2” platform parameter for EpiPath and ImmunoTwin cohorts and using the mLiftOver() function to convert the IDATS from the Naumova et al. study from the “EPIC” source platform to the “EPICv2” target platform. The openSesame() function was then used to perform data pre-processing with the “QCDPB” prep argument in the recommended human-specific preparation code. Quantile normalization was applied to the combined cohort data after background correction using signals from out-of-band probes and filtering out probes detected with p-values greater than 0.05.

The sva R package (version 3.54.0) was used for batch correction. To limit DNA methylation bias, the analysis excluded probes associated with single nucleotide polymorphisms (SNPs) and the X and Y chromosomes. The getBetas() function was used to create the β-values matrix and subsequently annotated using sample metadata and manifest file for the hg38 genome build (https://zwdzwd.github.io/InfiniumAnnotation#human/).

The sesame DML() function was used for differential methylation analysis with the contrast specified to the sample group. The covariates of age and sex were also included in this analysis. The summaryExtractTest() function was incorporated to generate the summary table with results showing and FDR < 0.05. To perform the partial least squares-discriminant analysis (PLS-DA), the mixOmics R package (version 6.30.0) was used and the clusterProfiler package (version 4.14.4) generated the hierarchical clustering, gene ontology and KEGG pathway analysis. Principal Component Analysis were performed using the general R stats package with prcomp() function (version 4.4.2). For data visualisation, metafor (version 4.8-0), ggplot2 (version 3.5.1), cowplot (version 1.1.3), corrplot (version 0.95), and ComplexHeatmap (version 2.22.0) were used for figure generation.

The polyepigenetic score used for analysis in R Studio (R version 4.4.2) with the R Base Package incorporates the formula for each individual (*i* = 1 to 227) by taking the sum of the beta value of each CpG (*c*) multiplied by its weighted value (*w*):

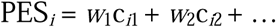

## Results

### DNA methylation analysis overview

The final analysis was performed on 227 participants from three cohorts (Table 1). This number included 129 controls and 78 adversity exposed individuals whovaried in timing of adversity exposure and age of study sampling. The analysis strategy highlights two approaches to examine the cohort data. First, individual cohorts were compared to identify common differentially methylated CpGs and genes. The second approach involved combining data from the 3 cohorts to perform an epigenome wide association study (EWAS) (Figure 1).

**Figure 1:**
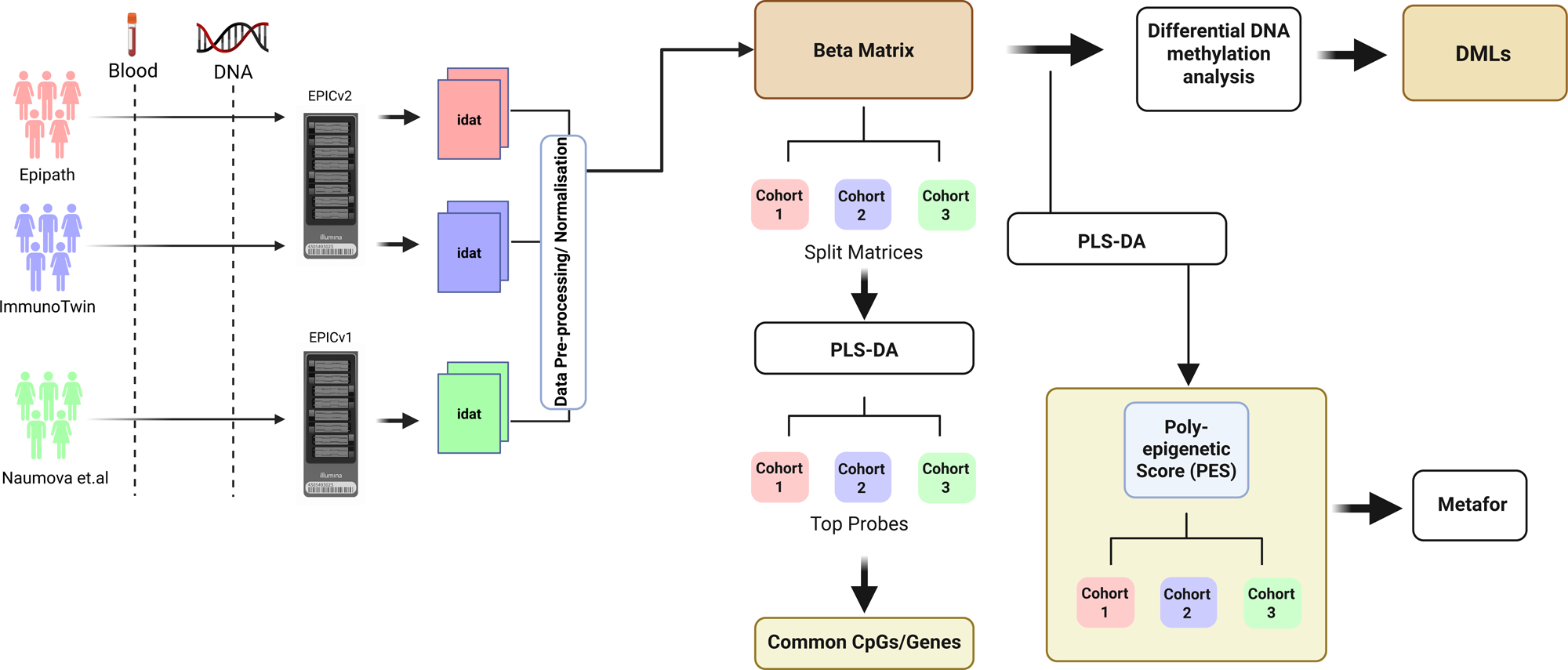
Outline of Analysis Strategy. The CpGs with the highest VIP score for each cohort were first identified to examine group differences and to identify common significant CpG sites and genes. An EWAS was then performed on the combined beta matrix to highlight differentially methylated loci and to calculate the poly-epigenetic score with the most significant CpGs and highest VIP score.

### Identification of unique and common biological signatures within the cohorts

To examine cohort differences, a first analysis was conducted on the methylation beta matrices of each cohort separately. Firstly, a partial least squares discriminant analysis (PLS-DA) was performed on each cohort’s corrected beta matrix to capture the differences between controls and ELA participants between studies (Supplementary Figure1A - C). The PLS-DA results were then used to find CpG sites of interest for further analysis by calculating the Variable Importance Projection (VIP) for each locus. An optimal cut-off for VIP scores was identified for each cohort by calculating the best corresponding AUC (Area Under the Curve) value within a ROC curve (Supplementary Figure1D - F).

Secondly, a Principal Component Analysis (PCA) was performed for each cohort using the identified CpGs from this VIP threshold (Figures 2A - C). The first principal component (PC1) for EpiPath accounted for 26.95% of the total variance and the second principal component (PC2) explained 4.69% from 799 CpG sites with a VIP cutoff of 3.1 (Figure 2A, Supplementary Figure 1D). ImmunoTwin incorporated 1561 CpGs with a VIP cutoff of 2.9.Results from the PCA analysis of the selected CpGs showed that PC1 explained 22.28% of the variance while PC2 explained 4.13% of the variance (Figure 2B). Lastly, the Naumova et al. study included 2811 CpGs from a VIP cut-off of 2.2 (Supplementary Figure 1E). PC1 accounted for 53.79% of the total variance and with 6.1% explained by PC2 (Figure 2C).

**Figure 2.**
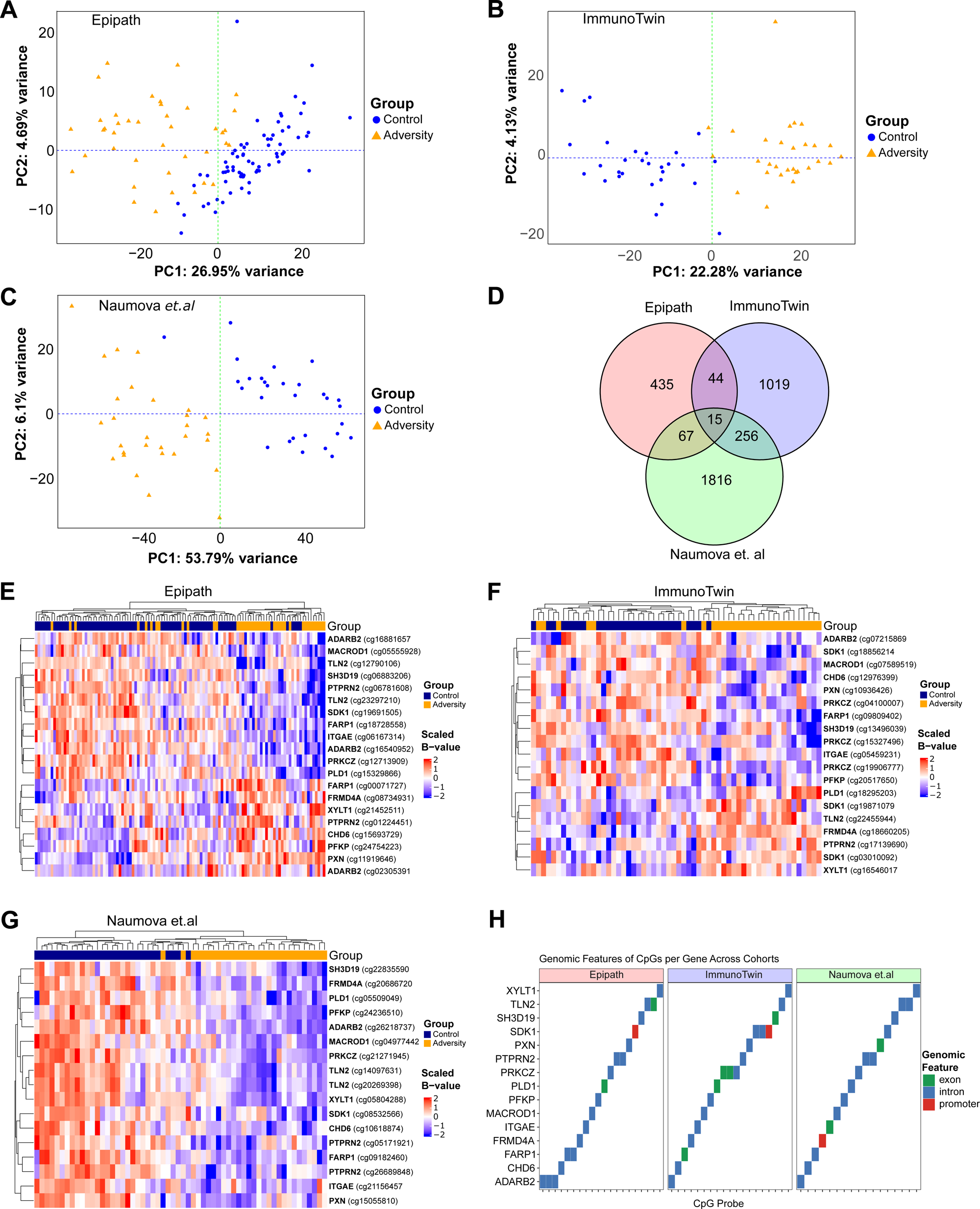
Principal Component Analyses (PCA) of VIP selected CpG loci across three cohorts. (A) PCA of the EpiPath cohort using 799 CpGs selected by a VIP cutoff of 3.1. (B) PCA of the ImmunoTwin cohort using 1561 CpGs (VIP cutoff 2.9). (C) PCA of the Naumova et al. cohort using 2811 CpGs (VIP cutoff 2.2) analysis of CpG associated genes across cohorts, identifying 15 common genes linked to neuronal development, chromatin remodeling, and metabolism. (E–G) Heatmaps showing methylation differences between controls and early life adversity participants in each cohort for the 15 shared genes. (H) Genomic feature distribution of the selected CpG sites across cohorts, showing localization within promoters, exons, and introns.

Next, the selected CpGs were compared across the 3 cohorts to identify common CpG loci that were differentially methylated. No CpG loci were found common to all 3 cohorts expect 7 CpG loci that were common in the ImmunoTwin and Naumova et.al studies (Supplementary Figure 1G). These 7 loci were associated with the genes, AFF3, CALML5, COL6A2, HR, KHNYN, MAB21L4 and AC137834.1 (associated with R3HDM2) (Supplemenatry Figure 1H – I).

Given that no CpGs were found common to all the 3 cohorts another analysis was carried out to identify CpG loci associated with common genes. 15 common genes linked to neuronal development, chromatin remodeling and metabolism were identified in all 3 cohorts (Figure 2D). Heatmaps were also generated to show differences in methylationbetween controls and adversity exposed participants in each of the cohorts for the 15 common genes(Figures 2E - G). Furthermore, an analysis to determine the genomic features associated with the identified CpG loci showed that across the cohorts CpGs were located in promoters (5.36%), exons (16.07%) and introns (78.57%)(Figure 2H).

### Exposure to adversity results in significant alterations of the epigenetic landscape

To investigate differences between the controls and adversity exposed individualsfrom all cohorts a second analysis was performed on the combined dataset. An epigenome-wide association study (EWAS) was performed with all 227 participants from each cohort. The analysis included age and sex as covariates and identified 18,600 differentially methylated CpG loci in the ELA group after correcting for multiple testing using the false discovery rate (FDR) (FDR < 0.05, Figure 3A). An increase in DNA methylation was observed in 1,480 CpGs associated with 1,091 genes while a decrease was observed in 17,120 sites associated with 7,602 genes (FDR < 0.05) (Figure 3B). Next, a more stringent FDR threshold (FDR < 0.01) was applied for further analysis to identify the most significant CpG sites and illustrated through a heatmap and a PLS-DA plot (Figure 3C-D). In the PLS-DA analysis, X-variate 1 accounted for 31% of the variance while X-variate 2 explained 3% of the variance.

**Figure 3.**
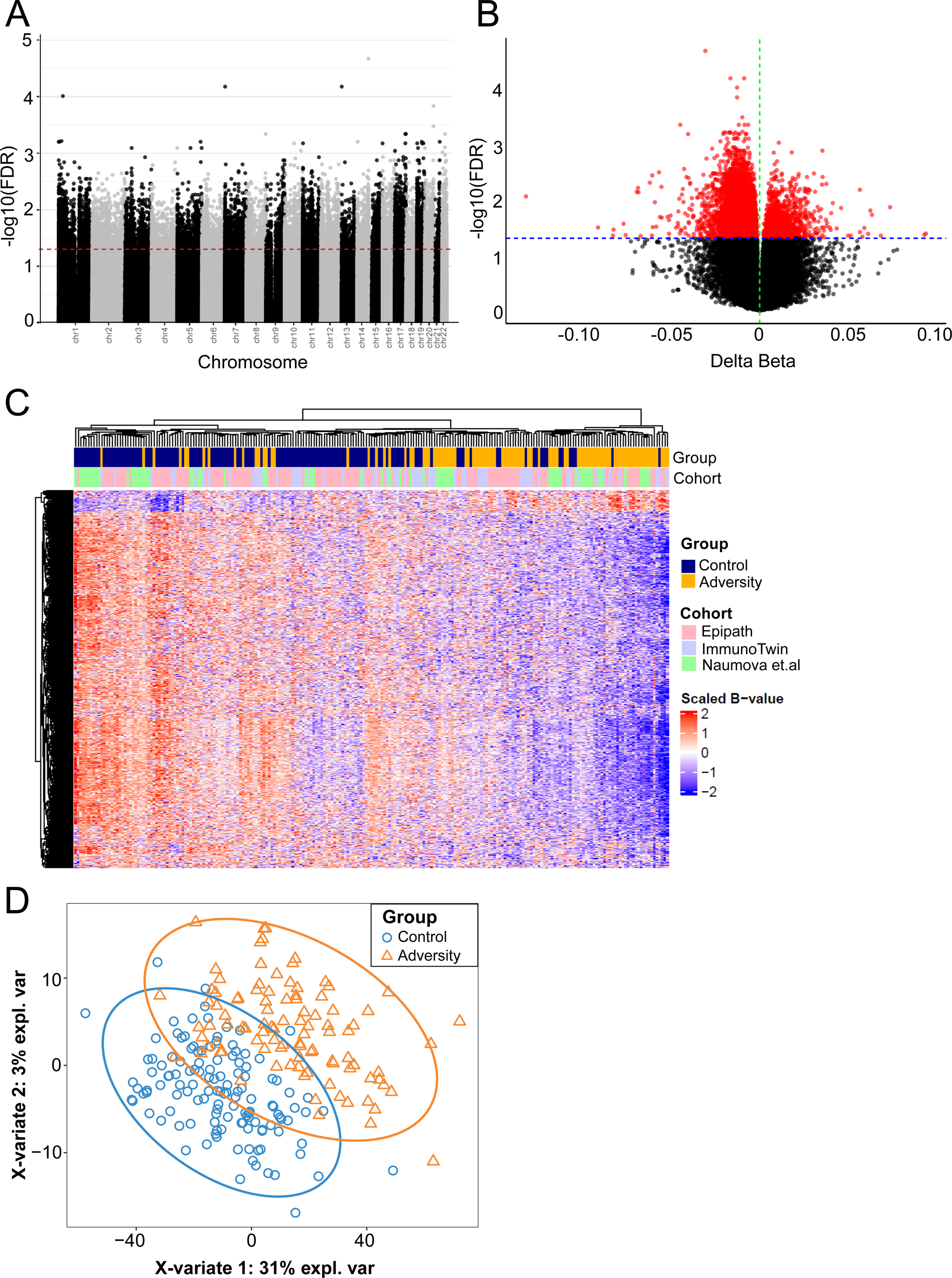
Epigenome-wide association analysis (EWAS) of combined cohorts. (A) EWAS performed on the combined dataset of 227 participants, adjusting for age and sex, identified 18,600 differentially methylated CpG loci associated with early-life adversity after FDR correction (FDR□<□0.05). (B) Distribution of methylation changes, showing hypermethylation in 1,480 CpGs (1,091 genes) and hypomethylation in 17,120 CpGs (7,602 genes). (C) Heatmap of the most significant CpG sites selected using a more stringent threshold (FDR□<□0.01). (D) Partial least squares discriminant analysis (PLS-DA) of the most significant CpGs.

The striking predominance of hypomethylation (>90% of significant sites) is consistent with the adaptive cost hypothesis, which predicts that adversity exposure triggers demethylation of genes involved in maintaining heightened physiological vigilance. Rather than reflecting stochastic degradation, this pattern may represent coordinated epigenetic reprogramming that “primes” stress-responsive, immune, and metabolic systems for anticipated environmental threats.

### Differential methylation in CpG loci of genes associated with psychosocial adversity

Adversity during the early-life to adolescent window is associated with differential gene expression potentially contributing to disease progression, including biological pathways linked to innate immune response [22] and pro-inflammatory function [23]. To explore pathway associations, a Gene Ontology (GO) and KEGG pathway enrichment analysis were both performed using the significant CpG sites (FDR < 0.05) to identify potential biological processes associated with early-life adversity (Figures 4A-B). Some of the significantly enriched pathways (p < 0.05) included oxytocin signalling, regulation of nervous system development, calcium signalling and insulin secretion. An additional KEGG analysis was performed using CpG sites with either an increase (Figure 4C) or decrease (Figure 4D) in methylation. These results revealed that genes associated with the oxytocin pathway, glutaminergic synapse, cortisol synthesis, insulin secretion and inflammatory mediator regulation of TRP channels had a global decrease in methylation. These findings support research that associates ELA with certain health risk factors such as autoimmune disorders and metabolic diseases like diabetes and obesity. In particular, the oxytocin system has been shown to be sensitive to early-life environments and potentially impacted by the level of parental care [24]. To further explore this pathway, a Concept NETwork (CNET) plot was created to examine connected pathways with genes associated with the oxytocin system within the three cohorts (Figure 5A).

**Figure 4.**
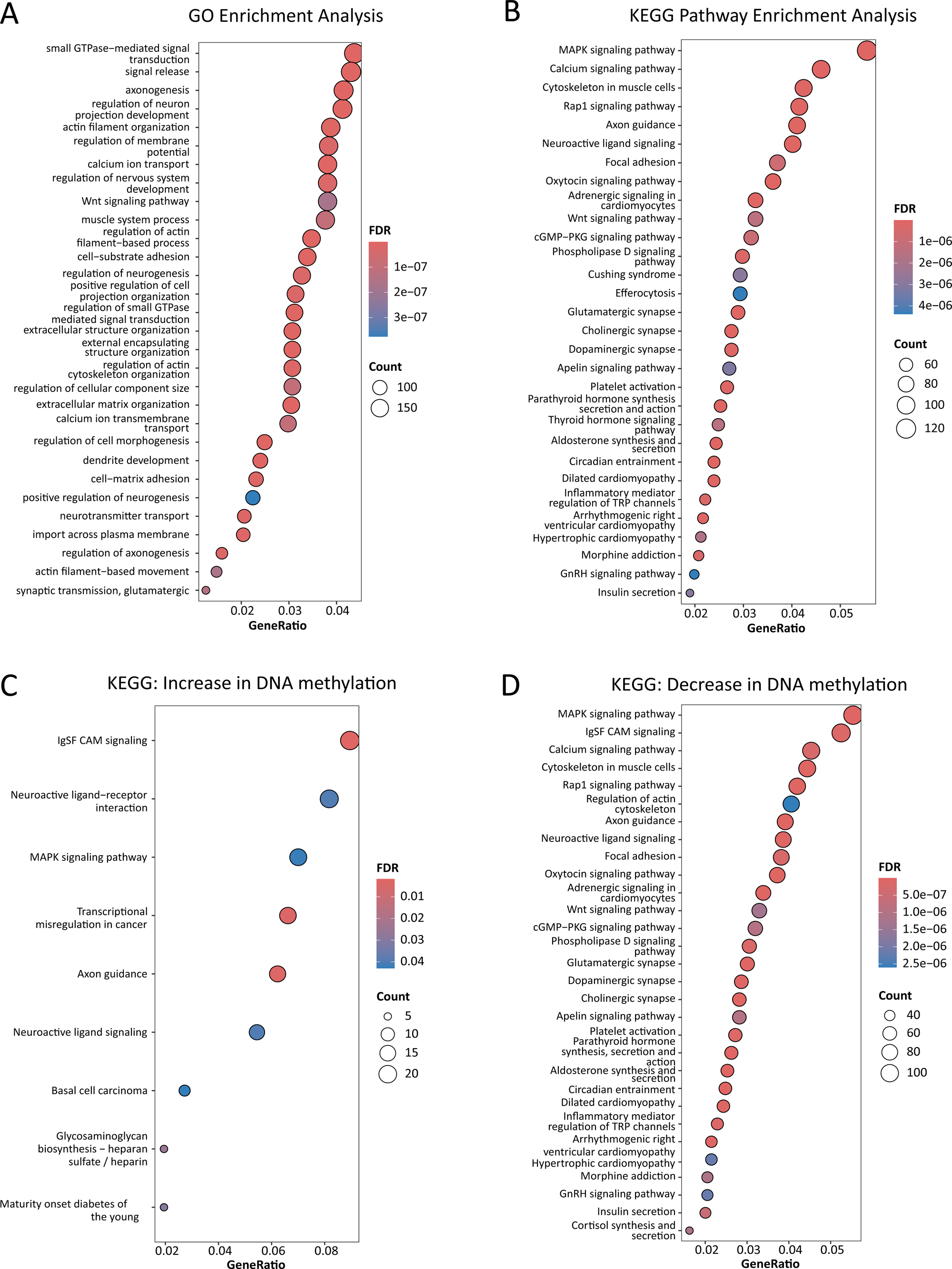
Gene Ontology and KEGG pathway enrichment analyses of differentially methylated CpG sites. (A–B) Gene Ontology (GO) and KEGG pathway enrichment analyses performed on significant CpG sites (FDR□<□0.05). (C–D) KEGG enrichment conducted separately for hypermethylated and hypomethylated CpG sites.

**Figure 5.**
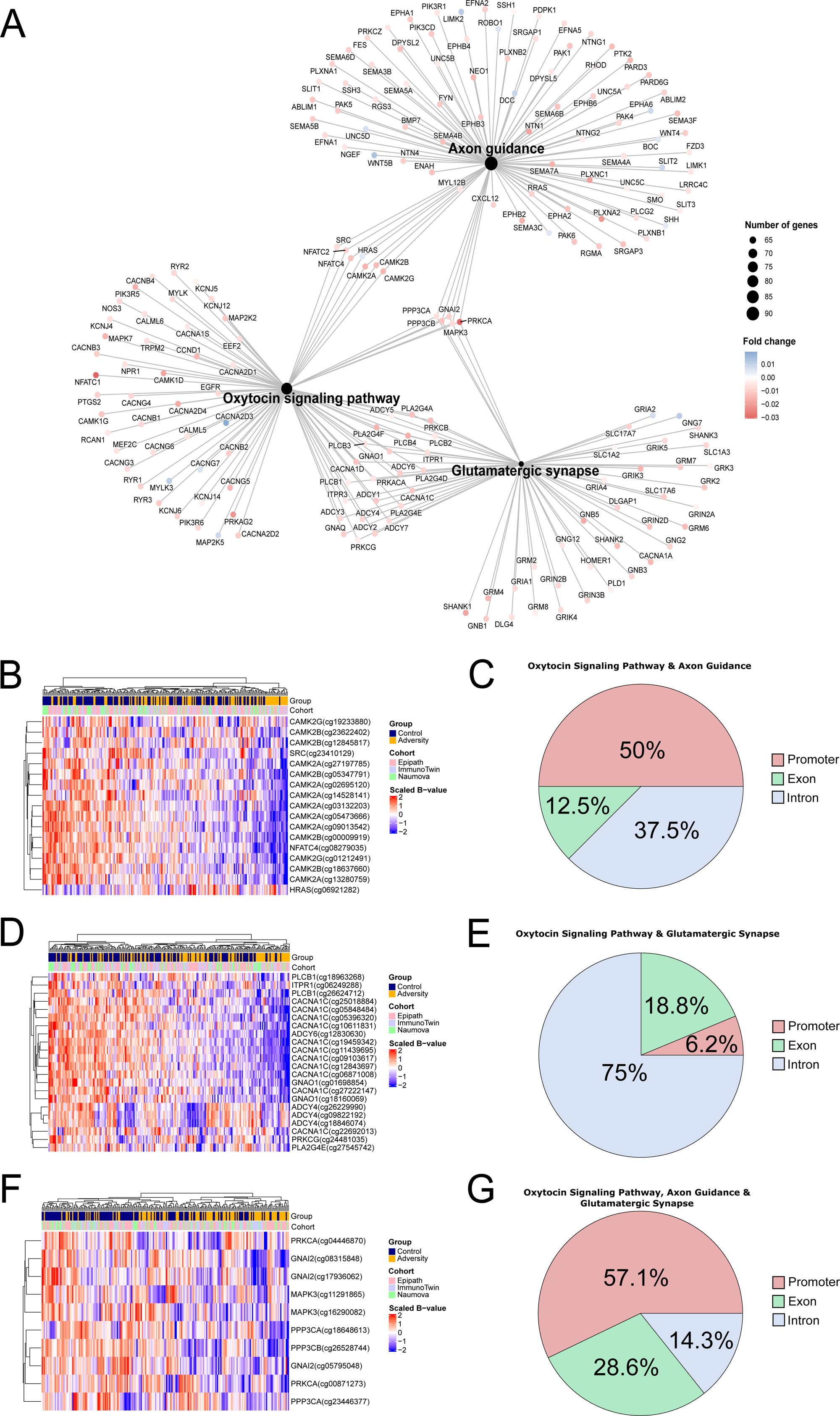
Oxytocin pathway–related network structure and genomic contexts of associated CpG loci. (A) CNET plot showing pathway–gene relationships for CpGs associated with oxytocin signaling across cohorts. (B–C) CpGs common to oxytocin signaling and axon guidance pathways. (D) CpGs overlapping between oxytocin signaling and glutamatergic synapse pathways. (E) Genomic feature distribution of CpG loci associated with oxytocin-related pathways. (F–G) Shared genes across oxytocin signaling, axon guidance, and glutamatergic synapse pathways and genomic annotation of their CpG sites.

Common genes between the oxytocin signaling pathway and axon guidance predominantly comprised of CpGs associated with 2 isoforms of Calcium/calmodulin[dependent protein kinase type II (CAMK2A and CAMK2B) and 50% were located in promoter regions (Figure 5B – C). On the other hand, common genes between oxytocin signaling and glutamatergic synapse comprised mostly of CpGs associated with CACNA1C, a gene that encodes an alpha-1 subunit of a voltage-dependent calcium channel (Figure 5D). Further analysis to determine the genomic features of identified CpG loci showed that 18.8% were located in exons, while a merger 6.2% were located in the promoter regions (Figure 5E). Lastly, the oxytocin signaling pathway, axon guidance and glutamatergic synapse pathways contained common genes, which included GNAI2, MAPK3, PPP3CA, PPP3CB and PRKCA (Figure 5F). 57.1% of CpG loci associated with these genes were found in promoters while 28.6% and 14.3% were located in exons and introns, respectively (Figure 5G).

### Poly-Epigenetic Score (PES) for adversity exposure

Polygenic methods have been widely used to develop tools such as genetic risk scores (GRS) as potential explanations of the variance within complex diseases and traits [25]. Recently, poly-epigenetic variations have been tested with similar methods to generate scores based on the weighted sum of beta values from pre-selected CpG sites [26]. Various methods for determining the appropriate weights and selected sites for scores have been used with formulations incorporating summary statistics from external studies or meta-analyses and p-value rankings of CpG sites following EWAS findings [27].

CpG probe selection for the poly-epigenetic score (PES) was performed using PLS-DA on the combined matrix of the three cohorts. The top CpG sites identified in X-variate 1 from the PLS-DA were also the most significant CpGs detected through the EWAS (FDR < 0.01) analysis of the combined matrix. The VIP scores for each CpG locus were ranked from highest to lowest and used as weights for the PES (Table 2).

To determine the number of CpG sites to include in the PES, the area under the curve (AUC) for Receiver Operating Characteristic (ROC) analysis was calculated to test different combinations of CpGs that had an FDR < 0.05 from the EWAS and high VIP score of X-variate 1. The AUC calculations identified200 CpGs were tested together (Figure x). As the AUC value remained relatively stable across the top CpG sites, the 200 CpGs with the highest VIP scores were chosen for the PES to maintain greater separation between scores and to allow for better comparisons of individuals.

### Lower PES scores in adversity-exposed individuals

To calculate the PES for each participant, the formula takes the sum of the VIP scores for each CpG multiplied by the corresponding beta values. A ROC curve analysis of the selected 200CpGs with the highest VIP scores showed an AUC value of 0.852(Figure 6A). Overall, low PES were observed in the adversity exposed group comparing the controls and adversity exposed individuals (Figures 6B – C). For the combined cohorts, adversity exposed individuals showed lower scores overall in comparison to the controls with significant group differences upon using Welch’s t-test (p < 2.2e-16) (Controls: mean PES +/- SD = 560.34 +/-5.10) (Adversity: mean PES +/- SD = 559.80 +/- 6.53)(Figure 6B). The mean difference between the PES of the adversity exposed group and controls showed ImmunoTwin averaging around five points below the controls while EpiPath and the Naumova et.al., study averaged around nine and thirteen points below controls, respectively (Figure 6D).

**Figure 6.**
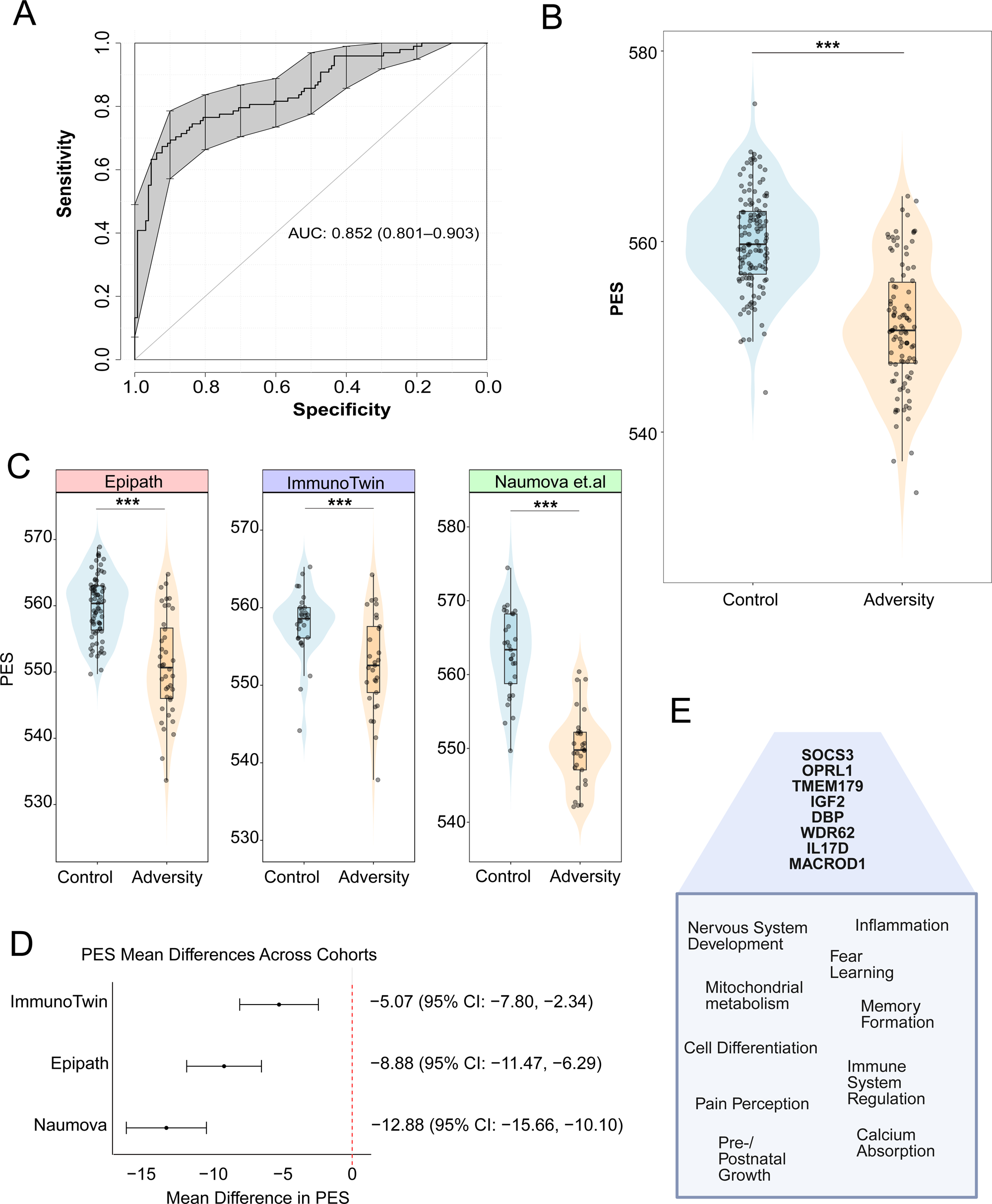
Polyepigenetic scores (PES) in adversity-exposed and control participants. (A) ROC curve analysis of the top 200 CpGs ranked by VIP score, showing an AUC of 0.852 for distinguishing adversity-exposed individuals from controls. (B) Polyepigenetic scores (PES), calculated as the sum of CpG-specific VIP values multiplied by corresponding beta values, were significantly lower in adversity-exposed individuals across the combined cohorts (Welch’s t-test, *p* < 2.2□×□10□¹□). (C–D) Cohort-specific PES differences, showing mean differences within each cohort relative to controls. (E) Top CpGs contributing to the PES included loci within genes previously linked to mental-health, metabolic, and mitochondrial responses to early-life adversity.

In addition, genes of interest linked to health outcomes of adversity exposures were identified in the top 10 CpG sites from the PES. These included CpGs found in the exon and shore region of the OPRL1 gene (chromosome 20) and in the CpG island region of the SOCS3 gene (chromosome 17). Recently, a study examining the OPRL1 gene found that moderate to high methylation had a moderating effect for depressive systems in individuals exposed to childhood trauma [28]. Researchers have also seen associations between obesity and hypomethylation of CpG sites within the SOCS3 gene [29]. The gene MACROD1, which encodes the mitochondrially localised mono-ADP ribosylhyrolase 1 protein, was present in the list of 15 common genes (Figure 2D – G) and was also found among the CpGs that made up the PES. (Figure 6E).

As the PES was developed using the Illumina EPIC and EPIC v2.0 arrays, a second PES was also calculated to allow for incorporation of DNA methylation studies previously published using the Illumina 450K BeadChip array system. A total of 70 CpGs out of the 200 CpGs from the first PES were found in the 450K arrays. These combined CpG sites showed similar AUC values when tested with the same scoring calculation.

### Decreased methylation in ELA for highest weighted CpGs in PES

To further explore the PES, the top 10 CpG sites with the highest VIP scores were further analysed with the metafor R package. As some heterogeneity was expected due to the participant differences and methods in each study, a random-effects model was performed to show the mean difference of methylation beta values between ELAs and Controls. Forest plots illustrate overall effect sizes in CpG sites for each study, demonstrating a decrease in methylation values for ELA participants (Figure 7A-D). Mean differences close to or above zero imply little or no difference in methylation levels between groups. Overall, larger differences in mean values were observed in the Naumova et al. study and EpiPath cohort. The CpGs with the highest VIP score weighting was cg14803282 and its associated gene is the transmembrane protein 179 (TMEM179) (Figure 7A). Although research on this protein’s function is limited, a previously published study has found it potentially plays and essential role in mitochondrial function, particularly in oligodendrocyte precursor cells that are later involved with the nervous system and myelination of neuronal axons [30]. The second highest CpG, cg04906043 had stronger differences between ELAs and Controls for the cohorts with institutionalisation exposure during early-life (Figure 7B). This CpG is associated with Interleukin-17D (IL17D) of the IL-17 cytokine family normally promoting host immune response, but a previous study using mouse models has differentiated IL17D as potentially reducing CD8 T cell functions during certain bacterial and viral infections [31]. Forest plots were also made for the same CpGs associated with SOCS3 and OPRL1 previously highlighted with box plots in Figure 6. While EpiPath and Naumova et al. study participants show larger methylation differences between groups for the SOCS3 linked CpG, cg11047325 (Figure 7C), visible effect sizes can be seen for all studies in the OPRL1 associated site cg15824741 (Figure 7D).

**Figure 7.**
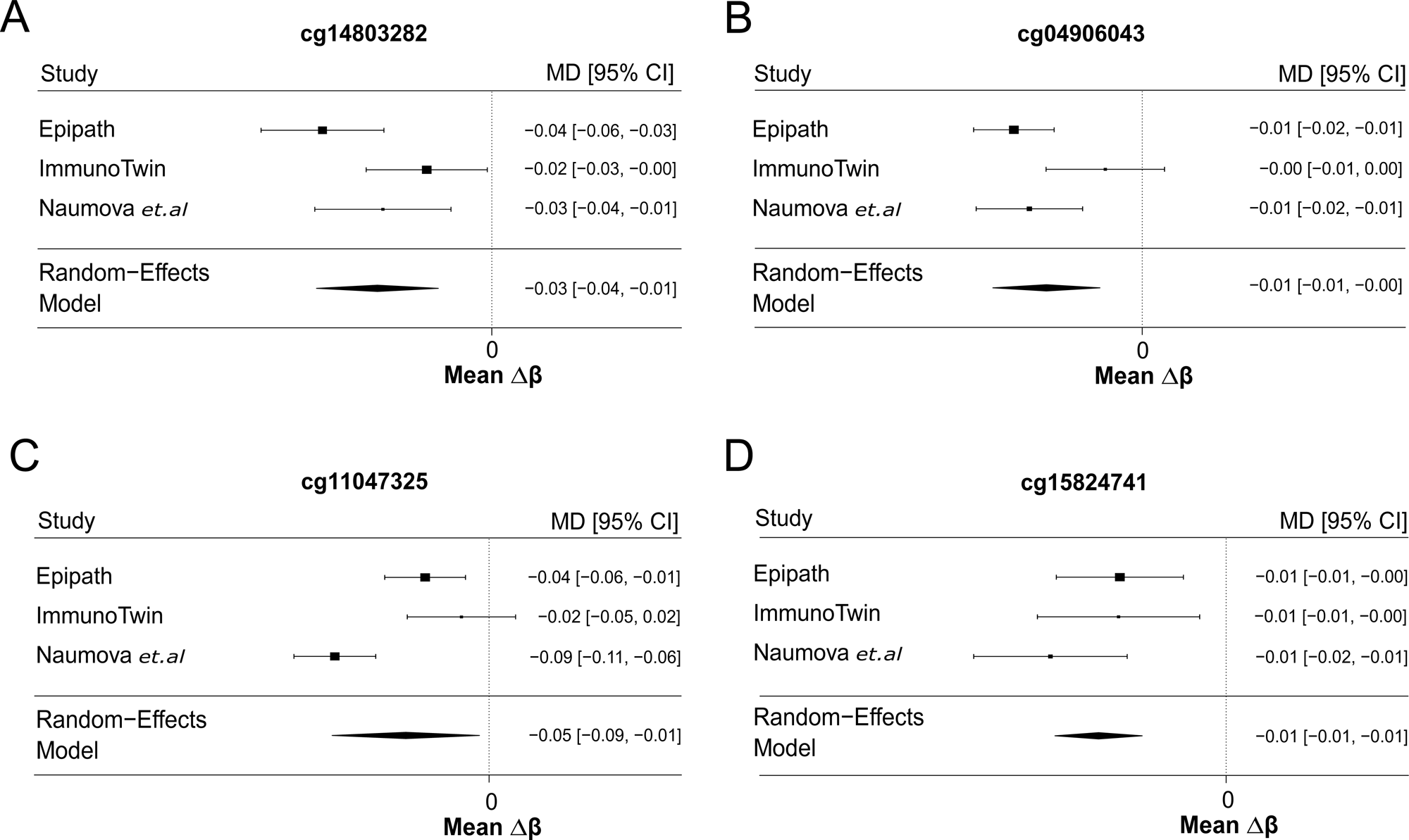
Forest plots of CpG-specific effect sizes across cohorts. (A–D) Forest plots showing effect sizes for top CpG sites across the three cohorts. Mean differences near or above zero indicate minimal methylation change between groups. (A) cg14803282, the CpG with the highest VIP score, associated with *TMEM179*. (B) cg04906043, associated with *IL17D*. (C) cg11047325, located within *SOCS3*. (D) cg15824741, associated with *OPRL1*.

## Discussion

The current study shows that the early stages of development is particularly sensitive to adversity exposure and is biologically embedded through epigenetic imprinting. Here CpG loci that have been associated with genes involved in biological processes such as inflammation, oxytocin signalling, and cortisol secretion were found to have persistent differential DNA methylation between controls and ELA individuals despite many years post-exposure. This study incorporated three cohorts with differences in age and type of adversity exposure. For EpiPath and the Naumova study participants, adversity in the form of deprivation in early childhood can have long-lasting effects on health trajectories from situations such as poverty, parental separation or loss, and inadequate social interaction [32]. Children from deprived backgrounds are also at risk of missing developmental milestones in non-cognitive skills like emotional stability and openness that help individuals become resilient in adulthood [33]. Exposure to adversity during the early developmental period is linked to later-life diagnosis of anxiety and depression disorders with severity of symptoms increasing alongside greater adverse experiences [34]. Following our EWAS, this study identified hypomethylation in CpG sites linked to SOCS3, which in addition to being associated with increased risk of obesity [29], previous research has found associations with early-life growth factors whereby lower methylation in exon regions of SOCS3 were associated with lower height and stunting risks in children [35]. For the ImmunoTwin cohort, the adolescent window has been shown to be an important timeframe for autonomy and social development. Negative experiences during this period have also been associated with potential increases in emotional reactivity and first onset of psychological disorders [36]. Early life stressors have been shown to influence immune function in adolescents with evidence of post-institutionalised youth having increased inflammation in relation to IL6 and CRP (C-reactive protein) levels [37]. However, the adolescent window may offer a recalibration timeframe whereby more favourable environmental conditions positively direct an individual’s long-term health trajectory [38]. Our study also found differential methylation in CpGs linked to OPRL1 expression, particularly decreased methylation in ELA participants, which has been linked to higher instances of psychosocial stress and unhealthy or risky behaviours such as binge drinking in adolescents [39].

The predominant hypomethylation observed in adversity-exposed individuals (>90% of differentially methylated sites) challenges traditional models of adversity as causing cumulative biological “damage.” Instead, our findings align with the adaptive cost hypothesis, which proposes that early-life adversity triggers adaptive epigenetic reprogramming designed to maintain heightened vigilance and responsivity to potential threats.

Under this framework, the widespread demethylation we observe in pathways involving oxytocin signalling, calcium dynamics, and nervous system development may reflect a coordinated biological strategy: maintaining these systems in a “primed” state ready to detect and respond to social and environmental stressors. This interpretation is supported by three key observations: Firstly, hypomethylation was particularly pronounced in genes involved in threat detection and social information processing (e.g., oxytocin pathway, CAMK2A/B, CACNA1C). In an unpredictable early environment, maintaining heightened sensitivity to social cues and potential threats may be adaptive, even if it incurs physiological costs. The oxytocin system, which showed significant hypomethylation across all cohorts, is central to social bonding and threat assessment [40, 41]—precisely the domains where adversity-exposed individuals must maintain vigilance. Next, the persistent nature of these methylation changes years after adversity exposure (particularly evident in our adult EpiPath cohort) suggests they represent stable recalibration rather than acute stress responses. This aligns with the adaptive cost model’s prediction that early environmental calibration creates lasting “biological priors” about threat probability, similar to Bayesian updating of internal models[15]. Finally, the cohort-specific differences in PES scores (largest effects in early institutional care cohorts, smaller effects in twins experiencing psychosocial stress) may reflect dose-dependent recalibration: more severe adversity necessitates more extensive epigenetic reprogramming to maintain adequate vigilance.

Critically, the adaptive cost framework reframes health disparities associated with early adversity not as inevitable consequences of “biological damage,” but as the physiological price of maintaining adaptive vigilance in environments perceived as threatening. This distinction has important implications for intervention: rather than attempting to “repair” damaged systems, effective approaches may need to provide sustained safety cues that permit biological recalibration and reduced vigilance costs.

Following the results for our GO and KEGG analyses, previous research has identified the oxytocin pathway as playing an important role in parental care, social behaviour and communication skills in adults [42]. As the oxytocin pathway was associated with decreased methylation in this study, previous research in stress response has also identified lower levels of methylation in the promoter of the OXTR gene for females exposed to stressful parenting styles during adolescence [43]. In addition, this study identified significantly enriched pathways involving calcium signalling and nervous system development. Early-life stressors may lead to disruptions in intracellular calcium processes involved in neurodevelopment, potentially increasing the risk factor for psychiatric disorders like schizophrenia [44]. Furthermore, calcium signalling has been identified as a possible pathway associated with the establishment of stress responsivity and sensitivity in the HPA-axis during early brain development [45]. Pathways impacting nervous system development have been shown to be affected by early-life adversity as chronic inflammation may alter emotional regulation, particularly leading to maladaptive strategies that can increase an individual’s perceived stress [46]. Interestingly, alterations in brain functions involving emotional processing have also been reported in post-institutionalised individuals who showed patterns of greater amygdala activation, potentially a beneficial adaptive response to stressful environments [47]. These findings highlight the importance of identifying children exposed to severe forms of early adversity like maternal separation to improve programs for early intervention.

Furthermore, our study showed common gene interactions between the oxytocin pathway, axon guidance and glutamatergic synapse pathways. Oxytocin signalling has been linked to axon guidance as being one pathway participating in the formation of hypothalamic axons required for later secretion of neuropeptides like oxytocin and arginine-vasopressin into the bloodstream through the neuroendocrine system [48]. In addition, A recently published study has found connections between transient oxytocin exposure and synaptogenesis in glutamatergic pyramidal neurons of the hippocampus in early neuronal formation, potentially influencing brain development and social behaviour through OXT maintenance of hippocampal excitatory and inhibitory neurons [49]. Two genes associated with the oxytocin pathway in our CNET plot were identified as HRAS and GNAI2 and were linked to CpGs with differential methylation patterns in the promoter regions. In a previously published study, hypomethylation in HRAS oncogenes potentially linked to cancer was found to have increasing cross-generational effects in children when one or more parents and grandparents suffered from alcohol dependence [50]. The HRAS oncogene has also been associated with developmental disorders such as Costello syndrome that is linked to abnormal growth and facial features and disrupted brain development [51]. Additionally, as one of the main G proteins, biological functions associated with GNAI2 include inflammation and hormonal regulation and signalling in hormones such as epinephrine and somatostatin [52]. Furthermore, metabolic functions within the central nervous system and adipose tissue have been linked to G-protein-coupled (GPCR)-signalling and dysregulation of this pathway is potentially involved with the development of obesity and diabetes [53]. The oxytocin pathway’s hypomethylation in adversity-exposed individuals is particularly noteworthy through an active cost lens. Oxytocin signalling regulates not only social bonding but also social threat detection and defensive responses[54]. Maintaining this system in a demethylated, transcriptionally accessible state may enable rapid deployment of both affiliative and defensive behaviors depending on social context—adaptive in unpredictable social environments but potentially costly in terms of sustained physiological activation. Our finding of differential methylation in CAMK2A/B and CACNA1C, key mediators of calcium signalling and synaptic plasticity, further supports the interpretation that adversity creates a “primed” neural substrate for processing social and threat-relevant information.

Polyepigenetic or methylation risk scores are commonly calculated through the weighted sums of DNA methylation beta values from CpG sites of interest [27]. The advancement of such scores has been aided by the growth in epigenome-wide association studies (EWAS) that identify CpG sites potentially associated with phenotypic traits being researched [26]. Polyepigenetic scores that use a combination of CpG sites may help explain a greater amount of variance in traits compared to individual CpG sites [55]. In this study, our PES was developed using three cohorts with identified social experiences of early adversity. The observed differences in methylation, specifically hypomethylation in ELA participants, within these CpG sites raise important questions about how adversity possibly impacts gene expression linked to negative environmental stressors. In addition, previous research has identified several CpG sites associated with persistent methylation changes past the postnatal period, particularly between birth and ten years of age [56]. The site cg11047325 from this list is included in our PES and is found in an island region of the SOCS3 gene. This site showed larger differences in methylation between ELAs and Controls for the EpiPath and Naumova et al. studies in the forest plot. Interestingly, three CpG sites associated with early childhood height in the previously mentioned study focused on SOCS3 [35] are also found in the top 200 CpGs of our PES, including cg11047325, cg13343932, and cg18181703. In addition, methylation in the site cg18181703 has been shown to be negatively correlated with obesity and certain traits of multiple metabolic syndrome (MetS) such as BMI and waist to height ratio [57].

Limitations to this study include the potential bias for population differences among our cohorts. Despite removing probes associated with SNPs during analysis, the generalisability of our findings may be impacted by these factors due to similarities shared among participants. In addition, although in EpiPath and Naumova et al. studies ELA participants both underwent early institutionalisation, the socio-economic differences between countries of residence could influence the overall level of adversity experienced by these groups. Furthermore, the ImmunoTwin cohort defines adversity as perceived social stress, potentially influencing the overall level of adversity being captured by the PES. As blood samples in our cohorts were collected at different ages, these factors may also play a role in the interpretation of our findings due to DNA methylation alterations that occur with increasing age [58]. Lastly, due to small sample sizes in our cohorts, the statistical power of our PES may be impacted in terms of its predictive capabilities as larger datasets in polygenic scores have shown higher accuracy [25].

## Conclusion

In conclusion, our study demonstrates that epigenetic similarities in individuals exposed to early-life adversity can be identified across multiple cohorts. These common signatures are shared among different ethnicities and age groups. Our findings led to the development of a polyepigentic score that captures early negative life experiences between three different cohorts. Furthermore, this score does not appear to be specific to the type of adversity experienced by an individual. Scores within the higher and lower ranges of the PES potentially represent a first step towards identifying individual differences within the groups. However, our findings challenge deterministic models of adversity as causing uniform biological damage. Instead, the predominant hypomethylation signature we observe across three distinct cohorts suggests coordinated epigenetic reprogramming consistent with the active cost hypothesis: adversity triggers adaptive recalibration of threat detection, social cognition, and immune systems to match environmental unpredictability. While this recalibration may be adaptive in threatening contexts, it incurs ongoing physiological costs that manifest as altered disease risk trajectories. The poly-epigenetic score we developed captures this shared signature of biological vigilance across diverse adversity exposures, providing a quantitative tool for assessing the extent of adversity-induced recalibration. Critically, framing these methylation changes as active cost rather than passive damage suggests interventions should focus on providing sustained environmental safety to permit biological recalibration, rather than attempting to reverse “damaged” systems.

## Data Availability

All data produced in the present study are available upon reasonable request to the authors

## Author Contributions

This manuscript was conceptualised by AM and JDT. For this element of the study: literature review: MB, JLC, JDT and AM ; data collection: AM, JLC SM ; data curation and analysis: MB, AM, JLC; visualisation: AM, MB; writing – original draft: MB and AM; scientific refinement – MB, JLC, JDT and AM; manuscript review and editing: all authors. All authors read and approved the final version of the manuscript.

## Acknowledgments

The authors would like to thank the individuals from each cohort for their participation in the studies. The authors would also like to express their appreciation to the researchers from the Naumova et al. study (Prof. Elena Grigorenko and Prof. Oxana Naumova) for providing their data for use in this current study. Further appreciation is given to Pauline Guébels for her technical support in the laboratory.

## Supplementary Material

Supplementary data for this article has been provided.

## Data Availability

Data is available upon request to the authors.

---

**Supplementary Figure 1.** Identification of discriminating CpG loci across cohorts. (A–C) Partial least squares discriminant analyses (PLS DA) performed separately for each cohort using corrected methylation beta matrices to distinguish controls from early life adversity participants. (D–F) Determination of optimal VIP score cutoffs for each cohort based on ROC curve derived AUC values. (G) Overlap of VIP selected CpG loci between cohorts, identifying shared CpGs between the ImmunoTwin and Naumova et al. cohorts. (H–I) Distribution of gene features of common CpG loci, corresponding to AFF3, CALML5, COL6A2, HR, KHNYN, MAB21L4, and AC137834.1 (associated with R3HDM2).

|  | *Variable* | *EpiPath* | *ImmunoTwin* | *Naumova et al.*<br>*study (2019)* |
| --- | --- | --- | --- | --- |
| *Demographics* | Controls | 73 | 29 | 29 |
|  | Adversity | 42 | 29 | 29 |
|  | Age | 20-25.5 (years) | 20-30 (years) | 8-35 (months) |
|  | Sex (Female) | 57% | 64% | 34% |
|  | Residence | Luxembourg and<br>Greater Region | Germany | Saint-Petersburg,<br>Russia |
|  | Adversity<br>Exposure | Institutionalisation<br>before adoption<br>(median 4.3 months) | Perceived stress | Institutionalisation |

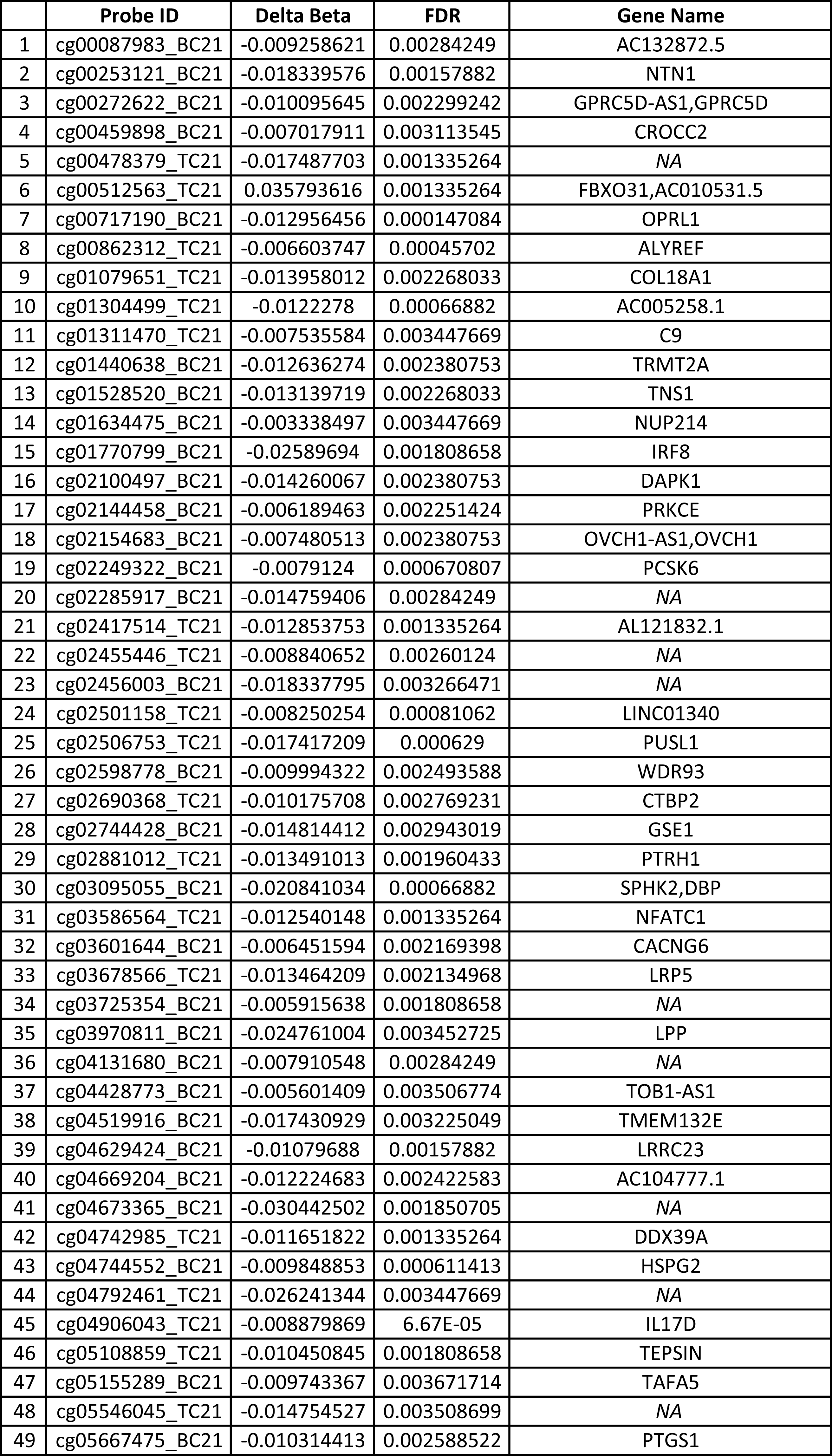
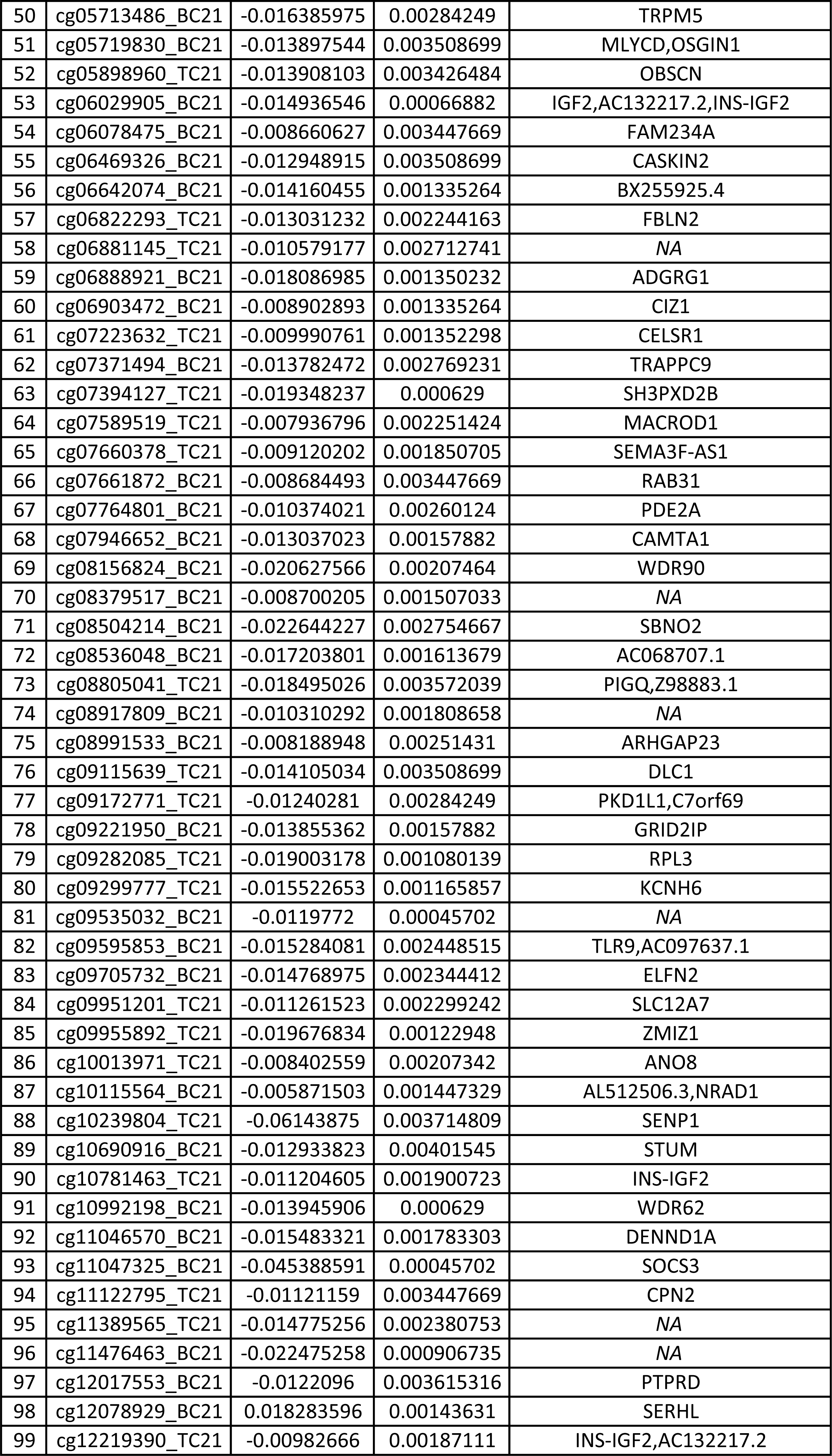
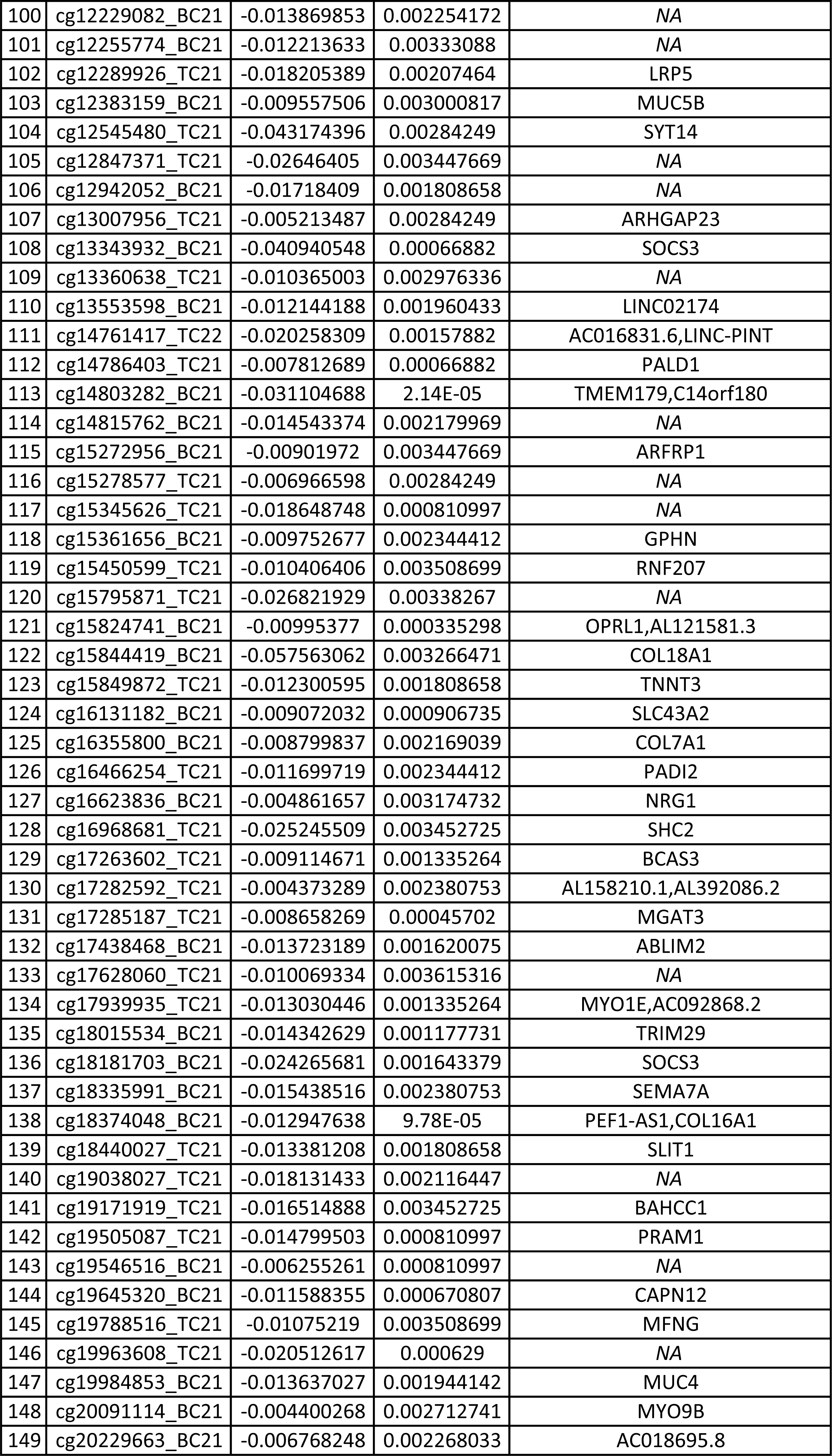
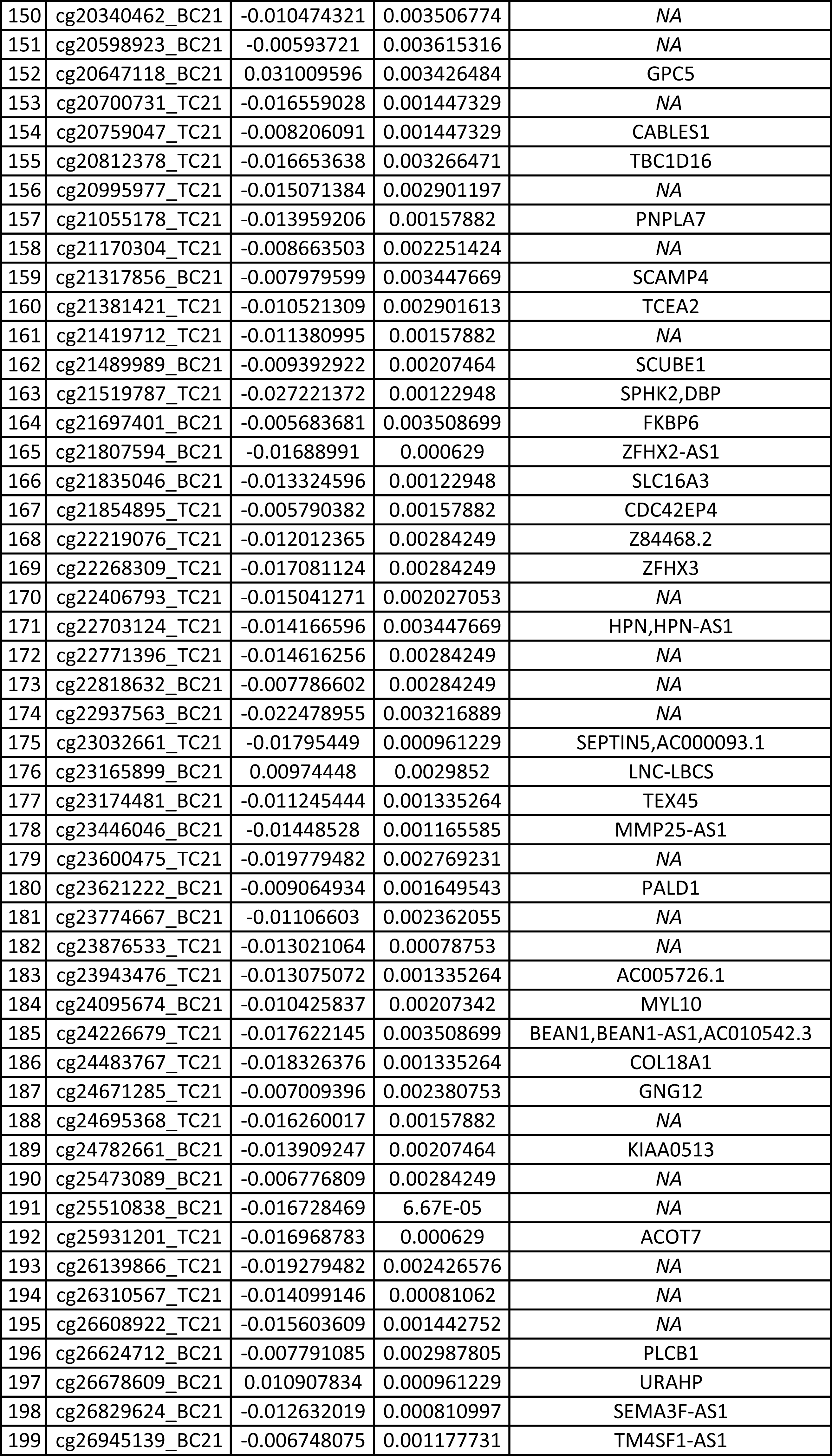
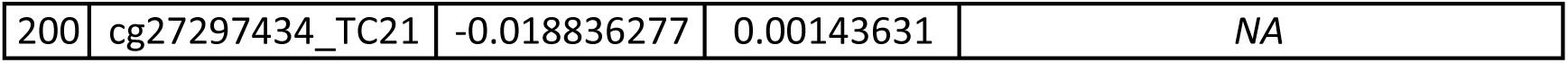

