## Supplementary figures and images for "Biological embedding of the pyscho-social environment; an Epigenetic Analysis of Adversity from Early-life to Adulthood"

### Supplementary Figure 1.pdf

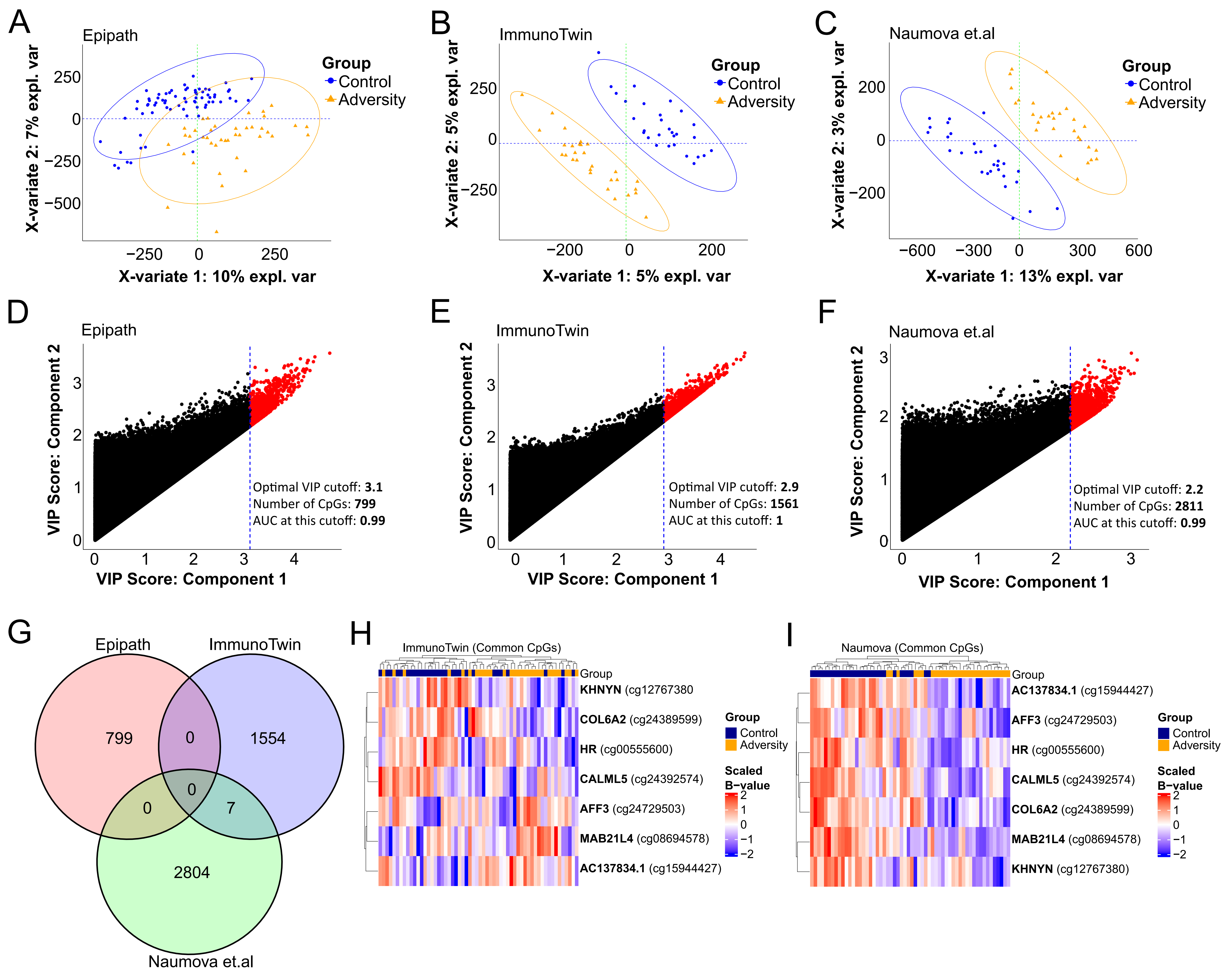
